# Mental health symptoms in a cohort of hospital healthcare workers following the first peak of the Covid-19 pandemic in the United Kingdom

**DOI:** 10.1101/2020.10.02.20205674

**Authors:** Kasun Wanigasooriya, Priyanka Palimar, David Naumann, Khalida Ismail, Jodie L. Fellows, Peter Logan, Christopher V. Thompson, Helen Bermingham, Andrew D. Beggs, Tariq Ismail

**Affiliations:** Institute of Biomedical Research, College of Medical and Dental Science, University of Birmingham, Vincent Drive, Edgbaston, Birmingham, B15 2TT; University Hospitals Birmingham NHS Foundation Trust, Mindelsohn Way, Edgbaston, Birmingham, B15 2WB; Department of Child and Adolescent Psychiatry, Forward Thinking Birmingham, Birmingham Women’s and Children’s Hospital NHS Foundation Trust, Birmingham B4 6NH; Department of Psychological Medicine, Institute of Psychiatry, Psychology and Neurosciences, King’s College London, Weston Education Centre, London, SE5 9RJ; Walsall Healthcare NHS Trust, Moat Road, Walsall, WS2 9PS; Sandwell and West Birmingham Hospitals NHS Trust, West Bromwich, B71 4HJ

**Keywords:** Anxiety, Depression, Post-Traumatic Stress Disorder (PTSD), Healthcare Workers, Covid-19, STAT-STRESS Covid-19 Study

## Abstract

**Background:** The Covid-19 pandemic is likely to lead to a significant increase in mental health disorders amongst healthcare workers (HCW).

**Aims:** We evaluated the prevalence of anxiety, depressive and post-traumatic stress disorder (PTSD) symptoms in a HCW population in the United Kingdom (UK), to identify subgroups most at risk.

**Methods:** An electronic survey was conducted between the 05/06/2020 and 31/07/2020 of all hospital HCW in the West Midlands, UK using clinically validated questionnaires: Patient Health Questionnaire-4 (PHQ-4) and the Impact of Event Scale-Revised (IES-R). Univariate analyses and adjusted logistic regression analyses were performed to estimate the strengths in associations.

**Results:** There were 2638 eligible participants who completed the survey (female: 79.5%, median age: 42 [IQR: 32-51] years). The prevalence rates of clinically significant symptoms of anxiety, depression and PTSD were 34.3%, 31.2% and 24.5% respectively. In adjusted analysis a history of mental health conditions was associated with clinically significant symptoms of anxiety (odds ratio 2.3 [95% CI 1.9–2.7]; *p*<0.001), depression (2.5 [2.1–3.0]; *p*<0.001) and PTSD (2.1 [1.7–2.5]; *p*<0.001). The availability of adequate personal protective equipment (PPE), wellbeing support and lower exposure to moral dilemmas at work demonstrated significant negative associations with former symptoms (*p*≤0.001).

**Conclusions:** We report a high prevalence of clinically significant symptoms of anxiety, depression and PTSD in hospital HCW following the initial Covid-19 pandemic peak in the UK. Those with a history of mental health conditions were most at risk. Adequate PPE availability, access to wellbeing support and reduced exposure to moral dilemmas may protect hospital HCW from mental health symptoms.

## Introduction

By September 2020, the Covid-19 pandemic caused by the novel SARS-CoV2 infection had claimed the lives of over 850,000 people worldwide.^1^ The pandemic has stretched the limits of healthcare systems to beyond capacity.^2^ Healthcare workers (HCW) have been exposed to numerous stressors and life events including: a rapid escalation in workload; sudden changes in roles and responsibilities including critical decision making (also referred to as moral injury);^3^ witnessing higher than the usual number of deaths; contracting the virus;^4,5^ caring for critically unwell family members; and grieving the loss of friends, family or colleagues. For many HCW, there has been a significant reduction in the usual sources of available social support due to changes in working schedules and social isolation measures.

Holmes et al. recently highlighted the importance of addressing the psychological impact of the pandemic on hospital staff as a key multidisciplinary mental health research priority.^6^ Several studies from Asia have evaluated the mental health impact of the Covid-19 pandemic on HCW.^7-11^ The consensus from these studies was that symptoms of mental health conditions were frequent amongst HCW during and after the peak of the pandemic in each respective country.^12^ This is akin to reports of increased anxiety, depression and post-traumatic stress disorder (PTSD) symptoms amongst HCW following the 2003 SARS outbreak in Asia.^13^ There are also concerns regarding the impact of PTSD symptoms on the National Health Service (NHS) workforce and the most effective interventions to support HCW.^6,14^

The mental health consequences of the Covid-19 pandemic on HCW in Western nations remains uncertain. The United Kingdom (UK) has the fifth highest number of deaths from Covid-19 worldwide.^1^ The peak of the pandemic in the UK occurred between March and May 2020.^15^ More than 100 HCW in the UK lost their lives to Covid-19.^5,16^ Several measures have been implemented to address the mental health sequelae of the pandemic, including the implementation of several staff wellbeing programmes and the allocation of over £5 million for mental health research.^17,18^ This study aims to describe the prevalence and risk factors for clinically significant symptoms of anxiety, depression and PTSD in a cohort of hospital based HCW from the UK, in the immediate aftermath of the Covid-19 pandemic peak.

## Methods

### Study design and setting

A cross-sectional survey of HCW employed in ten NHS acute general and mental health hospitals set in the West Midlands, UK was conducted between the 5^th^ of June 2020 and the 31^st^ of July 2020. The county has an ethnically and socio-economically diverse population.^19^ This region also observed a high incidence of Covid-19 cases and a high mortality rate.^20^ The study was approved by the UK Health Research Authority (HRA, Reference: 20/HRA/2865). Research Ethics Committee approval was not required for this study and this was confirmed by the HRA. Site specific approval was obtained from each of the research and development departments of all participating acute general (n=7) and mental health (n=3) NHS hospital Trusts. Informed consent was obtained from all participants and recorded electronically at the start of the study.

### Study participation

Eligible participants included all staff who worked or volunteered on-site at one of the participating hospitals for over 50% of their working week during the peak period of the Covid-19 pandemic in the UK. For this study, the latter was defined as the 23^rd^ of March 2020 to the 23^rd^ of May 2020. Staff who were on any type of leave for over 50% of the time during this period and those working from home or based in the community were not eligible to take part in this study. Eligible participants were invited to complete a confidential, voluntary electronic survey using the SurveyMonkey (San Mateo, California, USA) online survey administration and management platform. The survey was approximately 15 minutes long. It was distributed to participants via email, newsletters, posters, flyers and social media platforms to maximise reach and encourage participation.

### Exposure variables

The survey collected self-reported data on 24 independent variables. These included socio-demographic factors (age, gender, ethnicity, relationship status, number of dependents and immigration status); current health status (mental health conditions and physical illness); lifestyle factors (weekly smoking and alcohol consumption); employment factors (job title, total duration of employment in healthcare, the type of hospital, location of work within the hospital, whether infected patients were treated at the work place, patient-facing duties, availability of adequate personal protective equipment [PPE], availability and use of wellbeing support). Data were also collected on the impact of Covid-19 on professional life (redeployment, increased working hours, morally uncomfortable changes in the way they worked as a subjective measure of moral dilemma or injury); impact on personal life (diagnosis of Covid-19 in either self or a cohabitant, admission to hospital with Covid-19 in either self, close family or friend). The collected data were stored securely and processed confidentially in compliance with UK data protection law and the European Union General Data Protection Regulations.

### Mental health symptoms

The Patient Health Questionnaire-4 (PHQ-4) was used to assess symptoms of anxiety and depression. The Impact of Event Scale-Revised (IES-R) score was used to assess symptoms of PTSD. The psychometric properties of the PHQ-4 were acceptable as a screening tool. Studies reported a sensitivity of 86% and specificity of 83% for generalised anxiety disorder on the anxiety subscale for scores ≥3.^21^ The corresponding figures for the depression subscale were 83% and 90% respectively for major depressive disorder.^22^ The sensitivity and specificity of the IES-R were 91% and 82% respectively for a diagnosis of PTSD, where the cut-off score was ≥ 33.^23^ In this study, we used a score of ≥3 for each subscale of the PHQ-4, and ≥ 33 on the IES-R as the threshold scores to detect the presence of clinically significant symptoms of anxiety, depression and PTSD respectively.

### Data analysis

Data were collated using Microsoft Excel (Redmond, Washington, USA) and summarised as median (interquartile range) for non-normal data, and as proportions (percentage) for categorical data. Statistical analysis was performed using SPSS V.25 (IBM, New York, USA). The prevalence rates of mental health symptoms were calculated. For the categorical survey responses that were returned as “prefer not to say” or “unsure,” where combined analysis with the other responses was not possible, these were treated as missing data during statistical analysis. Univariate (unadjusted) analysis of measurements was performed for 24 predetermined, independent exposure variables using chi-square tests to assess the significance of association. Adjusted analysis using binary logistic regression modelling was performed to obtain odds ratios (OR) and 95% confidence intervals (95% CI). The regression model utilised all 24 measured independent variables to determine factors significantly associated with anxiety (PHQ-4 anxiety subscale ≥3), depressive (PHQ-4 depression subscale ≥3) or PTSD (IES-R≥33) symptoms. Multicollinearity of the 24 independent variables was assessed by calculating their variance inflation factors (VIFs). A *p-*value of less than 0.05 was assigned as the level of statistical significance.

## Results

A total of 2706 participants completed the survey of whom only 2638 met the eligibility criteria. Their median age was 42 (IQR: 32-51) years. The majority (n=2096; 79.5%) were female and 19.8% (n=524) were male (Table 1). Eighteen (n=18, <0.1%) respondents did not disclose their gender. Nearly a fifth (n=455, 17.2%) belonged to Black, Asian and Minority Ethnic (BAME) groups. The majority (n=2016, 76.4%) were in a relationship and nearly half (n=1298, 49.2%) had dependents. Whilst 84.1% (n=2216) of participants were UK residents since childhood, the remainder emigrated as adults. Around two fifths (n=983, 37.3%) reported a history of mental health conditions and 78.2% (n=769) were prescribed medication or psychological therapies. Approximately a quarter (n=667, 25.3%) reported a history of physical illness. Smokers accounted for 11.4% (n=301) of the sample and 65.4% (n=1725) consumed alcohol.

**Table 1:** Participant socio-demographic, lifestyle, health and employment factors. Percentages are demonstrated in rows; + Pearson chi-square statistical test used for univariate analysis to obtain p-values; * 2 ⨯ 2 chi-square analysis – excludes the “prefer not to say” group; * * 2 ⨯ 2 chi-square analysis of doctors and nurses (combined) versus other staff groups; PHQ-4 – Patient Health Questionnaire-4; PTSD – Post Traumatic Stress Disorder; BAME – Black, Asian and Minority Ethnic; PPE – Personal Protective Equipment. [i] Supplementary Table 1 provides a distribution of all participant job titles and locations of work, [ii] Includes participants based at acute mental health hospitals and those who were uncertain of the type of hospital, [iii] Includes those who reported no and those who were uncertain.

| | PHQ-4:<br>Anxiety Subscale $\geq 3$ | | | PHQ-4:<br>Depression Subscale $\geq 3$ | | | IES-R $\geq 33$<br>PTSD Symptoms: | | |
| --- | --- | --- | --- | --- | --- | --- | --- | --- | --- |
|  | Yes (%) | No (%) | *p*-value<sup>+</sup> | Yes (%) | No (%) | *p*-value<sup>+</sup> | Yes (%) | No (%) | *p*-value<sup>+</sup> |
| **Total** | 906 (34.3) | 1732 (65.7) | .. | 824 (31.2) | 1814 (68.8) | .. | 647 (24.5) | 1991 (75.5) | .. |
| **Age (years)** |  |  |  |  |  |  |  |  |  |
| $\leq 30$ | 241 (41.4) | 341 (58.6) | <0.001 | 209 (35.9) | 373 (64.1) | 0.001 | 161 (27.7) | 421 (72.3) | 0.013 |
| > 30 to 40 | 247 (37.1) | 418 (62.9) |  | 232 (34.9) | 433 (65.1) |  | 174 (26.2) | 491 (73.8) |  |
| > 40 to 50 | 215 (29.7) | 510 (70.3) |  | 199 (27.4) | 526 (72.6) |  | 169 (23.3) | 556 (76.7) |  |
| > 50 to 60 | 171 (29.8) | 403 (70.2) |  | 156 (27.2) | 418 (72.8) |  | 115 (20) | 459 (80) |  |
| > 60 | 32 (34.8) | 60 (65.2) |  | 28 (30.4) | 64 (69.6) |  | 28 (30.4) | 64 (69.6) |  |
| **Gender** |  |  |  |  |  |  |  |  |  |
| Female | 762 (36.4) | 1334 (63.6) | <0.001* | 685 (32.7) | 1411 (67.3) | <0.001* | 553 (26.4) | 1543 (73.6) | <0.001* |
| Male | 134 (25.6) | 390 (74.4) |  | 130 (24.8) | 394 (75.2) |  | 84 (16) | 440 (84) |  |
| Prefer not to say | 10 (55.6) | 8 (44.4) |  | 9 (50) | 9 (50) |  | 10 (55.6) | 8 (44.4) |  |
| **Ethnicity** |  |  |  |  |  |  |  |  |  |
| BAME | 154 (33.8) | 301 (66.2) | 0.806 | 135 (29.7) | 320 (70.3) | 0.428 | 129 (28.4) | 326 (71.6) | 0.037 |
| Caucasian | 752 (34.4) | 1431 (65.6) |  | 689 (31.6) | 1494 (68.4) |  | 518 (23.7) | 1665 (76.3) |  |
| **Relationship status** |  |  |  |  |  |  |  |  |  |
| In a relationship | 688 (34.1) | 1328 (65.9) | 0.672 | 601 (29.8) | 1415 (70.2) | 0.004 | 470 (23.3) | 1546 (76.7) | 0.009 |
| Not in a relationship | 218 (35) | 404 (65) |  | 223 (35.9) | 399 (64.1) |  | 177 (28.5) | 445 (71.5) |  |
| **Dependents** |  |  |  |  |  |  |  |  |  |
| Yes | 420 (32.4) | 878 (67.6) | 0.034 | 375 (28.9) | 923 (71.1) | 0.011 | 317 (24.4) | 981 (75.6) | 0.903 |
| No | 486 (36.3) | 854 (63.7) |  | 449 (33.5) | 891 (66.5) |  | 330 (24.6) | 1010 (75.4) |  |
| **Immigration status** |  |  |  |  |  |  |  |  |  |
| Emigrated to the UK | 128 (30.3) | 294 (69.7) | 0.058 | 115 (27.3) | 307 (72.7) | 0.054 | 109 (25.8) | 313 (74.2) | 0.497 |
| UK resident from childhood | 778 (35.1) | 1438 (64.9) |  | 709 (32) | 1507 (68) |  | 538 (24.3) | 1678 (75.7) |  |
| **History of mental health conditions** |  |  |  |  |  |  |  |  |  |
| Yes | 457 (46.5) | 526 (53.5) | <0.001* | 440 (44.8) | 543 (55.2) | <0.0001* | 327 (33.3) | 656 (66.7) | <0.001* |
| No | 418 (26.3) | 1172 (73.7) |  | 355 (22.3) | 1235 (77.7) |  | 294 (18.5) | 1296 (81.5) |  |
| Prefer not to say | 31 (47.7) | 34 (52.3) |  | 29 (44.6) | 36 (55.4) |  | 26 (40) | 39 (60) |  |
| **History of physical illness** |  |  |  |  |  |  |  |  |  |
| Yes | 252 (37.7) | 417 (62.3) | 0.018* | 245 (36.6) | 424 (63.4) | <0.001* | 200 (29.9) | 469 (70.1) | <0.001* |
| No | 632 (32.7) | 1303 (67.3) |  | 561 (29) | 1374 (71) |  | 432 (22.3) | 1503 (77.7) |  |
| Prefer not to say | 22 (64.7) | 12 (35.3) |  | 18 (52.9) | 16 (47.1) |  | 15 (44.1) | 19 (55.9) |  |
| **Smoker** |  |  |  |  |  |  |  |  |  |
| Yes | 131 (43.5) | 170 (56.5) | <0.001 | 132 (43.9) | 169 (56.1) | <0.001 | 106 (35.2) | 195 (64.8) | <0.001 |
| No | 775 (33.2) | 1562 (66.8) |  | 692 (29.6) | 1645 (70.4) |  | 541 (23.1) | 1796 (76.9) |  |
| **Alcohol consumer** |  |  |  |  |  |  |  |  |  |
| Yes | 561 (32.5) | 1164 (67.5) | 0.007 | 494 (28.6) | 1231 (71.4) | <0.001 | 377 (21.9) | 1348 (78.1) | <0.001 |
| No | 345 (37.8) | 568 (62.2) |  | 330 (36.1) | 583 (63.9) |  | 270 (29.6) | 643 (70.4) |  |
| **Job title** |  |  |  |  |  |  |  |  |  |
| Doctors | 101 (22) | 359 (78) | <0.001** | 82 (17.8) | 378 (82.2) | <0.001** | 61 (13.3) | 399 (86.7) | 0.149** |
| Nurses | 279 (36) | 496 (64) |  | 255 (32.9) | 520 (67.1) |  | 226 (29.2) | 549 (70.8) |  |
| Other<sup>i</sup> | 526 (37.5) | 877 (62.5) |  | 487 (34.7) | 916 (65.3) |  | 360 (25.7) | 1043 (74.3) |  |
| **Duration of employment within healthcare** |  |  |  |  |  |  |  |  |  |
| $\leq 10$ years | 360 (38.2) | 582 (61.8) | 0.002 | 350 (37.2) | 592 (62.8) | <0.001 | 270 (28.7) | 672 (71.3) | <0.001 |
| >10 years | 546 (32.2) | 1150 (67.8) |  | 474 (27.9) | 1222 (72.1) |  | 377 (22.2) | 1319 (77.8) |  |
| **Based at an acute general hospital** |  |  |  |  |  |  |  |  |  |
| Yes | 854 (34.2) | 1646 (65.8) | 0.396 | 786 (31.4) | 1714 (68.6) | 0.335 | 611 (24.4) | 1889 (75.6) | 0.662 |
| No<sup>ii</sup> | 52 (37.7) | 86 (62.3) |  | 38 (27.5) | 100 (72.5) |  | 36 (26.1) | 102 (73.9) |  |
| ***Location within hospital*** |  |  |  |  |  |  |  |  |  |
| *Inpatient Wards / ED / ITU* | 435 (36) | 774 (64) | 0.104 | 379 (31.3) | 830 (68.7) | 0.909 | 355 (29.4) | 854 (70.6) | <0.001 |
| *Other<sup>i</sup>* | 471 (33) | 958 (67) |  | 445 (31.1) | 984 (68.9) |  | 292 (20.4) | 1137 (79.6) |  |
| ***Hospital treated Covid-19 patients*** |  |  |  |  |  |  |  |  |  |
| *Yes* | 872 (34.2) | 1679 (65.8) | 0.344 | 793 (31.1) | 1758 (68.9) | 0.368 | 630 (24.7) | 1921 (75.3) | 0.272 |
| *No<sup>iii</sup>* | 34 (39.1) | 53 (60.9) |  | 31 (35.6) | 56 (64.4) |  | 17 (19.5) | 70 (80.5) |  |
| ***Patient-facing duties (i.e. frontline clinical staff)*** |  |  |  |  |  |  |  |  |  |
| *Yes* | 554 (35.6) | 1000 (64.4) | 0.091 | 465 (29.9) | 1089 (70.1) | 0.081 | 420 (27) | 1134 (73) | <0.001 |
| *No* | 352 (32.5) | 732 (67.5) |  | 359 (33.1) | 725 (66.9) |  | 227 (20.9) | 857 (79.1) |  |

Most respondents were nurses (n=775, 29.5%), followed by doctors (n=460, 17.4%). Many staff (n=1403, 53.1%) performing various other roles within the hospitals also took part in the survey (supplementary Table 1). A third of participants (n=942, 35.7%) had worked in healthcare for ten years or less. The majority worked in acute general hospitals (n=2500, 94.7%) and the remainder at mental health hospitals (n=91, 3.5%). A minority (n=47, 1.8%) were uncertain of the type of their hospital. Staff were based in inpatient wards (n=704, 26.7%), intensive therapy units (ITU, n=382, 14.5%), emergency departments (ED, n=123, 4.7%) and other locations within their respective hospitals (n=1429, 54.2%; see supplementary Table 1). The majority of participants (n=1554, 59%) reported that they undertook patient-facing duties (i.e. frontline clinical staff).

### Participant experiences

Just over a half (n=1452, 55%) reported that adequate PPE was available at their workplace (Table 2). The remainder reported that this was not the case (n=812, 30.8%), were unsure (n=322, 12.2%) or preferred not to comment (n=52, 2%). The majority were aware of wellbeing measures implemented by their employer (n=2064, 78.2%) but only 15.3% (n=405) accessed any form of psychological support during the study period. Thirty per cent (n=790) of participants were redeployed as a result of the pandemic and 38.5% (n=1015) reported increased working hours. Forty-five per cent (n=1208) reported morally uncomfortable changes in the way they worked. Of those who reported a diagnosis of Covid-19 (n=720, 27.3%), this was either confirmed on polymerase chain reaction testing of nasopharyngeal swabs (n=277), diagnosed by a clinician (n=47) or self-diagnosed based on symptoms and Public Health England guidance (n=396). Approximately a fifth of staff (n=522, 19.8%) also reported that a cohabitant had developed Covid-19 during the period in question. Only 17.1% (n=452) reported an admission to hospital with Covid-19 in self, a close family member or friend.

**Table 2:** The impact of the Covid-19 pandemic on participants. Percentages are demonstrated in rows; + Pearson chi-square statistical test used for univariate analysis to obtain p-values; PHQ-4 – Patient Health Questionnaire-4; PTSD – Post Traumatic Stress Disorder; BAME – Black, Asian and Minority Ethnic; PPE – Personal Protective Equipment. [i] Includes all forms of redeployment e.g. different speciality, department or hospital, [ii] Includes no and not applicable, [iii] Includes those that reported a decrease, no change and not applicable, [iv] Includes those that reported yes and prefer not to say, [v] Includes all participants who had symptomatic Covid-19 illness diagnosed by polymerase chain reaction testing, clinician diagnosed or self-diagnosed as per Public Health England guidance, [vi] Close family member – nuclear family, first degree relative whom the respondent lived with or associated on at least once a week; a close friend: A friend whom the participant lived with or associated with at least once a week.

| | PHQ-4:<br>Anxiety Subscale $\geq 3$ | | | PHQ-4:<br>Depression Subscale $\geq 3$ | | | IES-R $\geq 33$<br>PTSD Symptoms: | | |
| --- | --- | --- | --- | --- | --- | --- | --- | --- | --- |
|  | Yes (%) | No (%) | p-value<sup>+</sup> | Yes (%) | No (%) | p-value<sup>+</sup> | Yes (%) | No (%) | p-value<sup>+</sup> |
| **Total** | 906 (34.3) | 1732 (65.7) | .. | 824 (31.2) | 1814 (68.8) | .. | 647 (24.5) | 1991 (75.5) | .. |
| **Redeployment** |  |  |  |  |  |  |  |  |  |
| Reported redeployment<sup>i</sup> | 320 (36.7) | 553 (63.3) | 0.079 | 276 (31.6) | 597 (68.4) | 0.767 | 274 (31.4) | 599 (68.6) | <0.001 |
| Did not report redeployment<sup>ii</sup> | 586 (33.2) | 1179 (66.8) |  | 548 (31) | 1217 (69) |  | 373 (21.1) | 1392 (78.9) |  |
| **Increased working hours** |  |  |  |  |  |  |  |  |  |
| Reported increase | 348 (34.3) | 667 (65.7) | 0.96 | 331 (32.6) | 684 (67.4) | 0.228 | 273 (26.9) | 742 (73.1) | 0.025 |
| Did not report increase<sup>iii</sup> | 558 (34.4) | 1065 (65.6) |  | 493 (30.4) | 1130 (69.6) |  | 374 (23) | 1249 (77) |  |
| **Morally uncomfortable changes in the way they worked (moral dilemmas)** |  |  |  |  |  |  |  |  |  |
| Denied experiencing | 344 (25.5) | 1007 (74.5) | <0.001 | 337 (24.9) | 1014 (75.1) | <0.001 | 193 (14.3) | 1158 (85.7) | <0.001 |
| Did not deny experiencing<sup>iv</sup> | 562 (43.7) | 725 (56.3) |  | 487 (37.8) | 800 (62.2) |  | 454 (35.3) | 833 (64.7) |  |
| **Developed Covid-19** |  |  |  |  |  |  |  |  |  |
| Yes<sup>v</sup> | 262 (36.4) | 458 (63.6) | 0.175 | 238 (33.1) | 482 (66.9) | 0.217 | 209 (29) | 511 (71) | 0.001 |
| No | 644 (33.6) | 1274 (66.4) |  | 586 (30.6) | 1332 (69.4) |  | 438 (22.8) | 1480 (77.2) |  |
| **A cohabitant developed Covid-19** |  |  |  |  |  |  |  |  |  |
| Yes | 179 (34.3) | 343 (65.7) | 0.977 | 157 (30.1) | 365 (69.9) | 0.523 | 131 (25.1) | 391 (74.9) | 0.736 |
| No | 727 (34.4) | 1389 (65.6) |  | 667 (31.5) | 1449 (68.5) |  | 516 (24.4) | 1600 (75.6) |  |
| **Admission to hospital with Covid-19** |  |  |  |  |  |  |  |  |  |
| Self, close family or friend<sup>vi</sup> | 189 (41.8) | 263 (58.2) | <0.001 | 154 (34.1) | 298 (65.9) | 0.153 | 161 (35.6) | 291 (64.4) | <0.001 |
| No | 717 (32.8) | 1469 (67.2) |  | 670 (30.6) | 1516 (69.4) |  | 486 (22.2) | 1700 (77.8) |  |

### Significant mental health symptoms

On the anxiety subscale of the PHQ-4, 34.3% (n=906) scored ≥3; on the depression subscale of the PHQ-4, 31.2% (n=824) scored ≥3 and 24.5% (n=647) scored ≥33 on the IES-R (supplementary Fig. 1).

### Univariate analysis

Female participants, those with a history of mental health or physical conditions, were smokers or consumed alcohol, worked as a doctor or a nurse, and had worked for ten years or less, were significantly more likely to have higher prevalence rates of anxiety symptoms (Table 1). These independent variables were also significantly associated with higher rates of depressive and PTSD symptoms. On the other hand, participants who had adequate PPE and wellbeing support available, and did not experience morally uncomfortable changes in the way they worked, reported significantly lower rates of all mental health symptoms (Table 2). Of note, being admitted or having a close family member or friend being admitted with Covid-19 infection was associated with increased anxiety and PTSD symptoms but not depressive symptoms (Table 2).

### Logistic regression analysis

The adjusted logistic regression analysis was performed for 2534 participants (excluding any missing data) for each of the dependent variables: anxiety, depression and PTSD symptoms using the clinically significant thresholds previously stated. All 24 independent variables considered in the univariate analysis were included in the regression analysis. An assessment of multicollinearity of the 24 independent variables revealed all the VIFs to be less than two (maximum 1.6), indicating the validity of including all these independent variables in the logistic regression modelling.

#### i. Anxiety symptoms

The statistically significant associations were as follows: younger participants, females and those reporting an admission to hospital with Covid-19 in self, close family member or friend were around 50% more likely to report anxiety symptoms (*p*≤0.001) (Table 3). A history of mental health conditions was also significantly associated with a greater than two-fold odds of clinically significant anxiety symptoms (OR 2.3 [1.9-2.7]; *p*<0.001). Doctors and nurses were 20% less likely to report anxiety compared to other hospital HCW (OR 0.8 [0.6-0.9]; *p=*0.009). Furthermore, those who reported adequate PPE availability, wellbeing support availability and where no morally uncomfortable changes took place, were around 50% less likely to have anxiety symptoms (*p*<0.001; see Table 3 and supplementary Fig. 2). There were no significant associations between the remaining exposures and anxiety symptoms (supplementary Table 2).

**Table 3:** Factors associated with significant mental health symptoms on adjusted analysis. * Admission to hospital in self, close family or friend 2534 participants included in the analysis excluding 104 who were treated as missing data where responses were ambiguous (i.e. prefer not to say) and could not therefore be combined or analysed as separate categories. A total of 24 independent variables (as displayed on Table 1 and Table 2) were used in the analysis. Only the statistically significant (*p*<0.05) results are displayed on Table 3. Please refer to supplementary Table 2 for a full list of variables included in the regression analysis.

|  | **PHQ-4:**<br>**Anxiety Subscale**<br>**Score $\geq 3$** | **PHQ-4:**<br>**Depression Subscale**<br>**Score $\geq 3$** | **PTSD Symptoms:**<br>**IES-R Score $\geq 33$** |
| --- | --- | --- | --- |
| Age $\leq 40$ | 1.4 (1.1 – 1.7); $p=0.001$ | -- | - |
| Female gender | 1.5 (1.2 – 1.9); $p=0.001$ | - | 1.7 (1.3 – 2.2); $p<0.001$ |
| A history of mental health conditions | 2.3 (1.9 – 2.7); $p<0.001$ | 2.5 (2.1 – 3.0); $p<0.001$ | 2.1 (1.7 – 2.5); $p<0.001$ |
| A history of physical illness | - | - | 1.4 (1.1 – 1.7); $p=0.009$ |
| Smoker | - | 1.5 (1.1 – 1.9); $p=0.005$ | 1.5 (1.1 – 1.9); $p=0.012$ |
| Alcohol consumer | - | 0.8 (0.7-0.9); $p=0.028$ | 0.8 (0.6 – 0.9); $p=0.038$ |
| Worked as a doctor or nurse | 0.8 (0.6-0.9); $p=0.009$ | - | 0.8 (0.6 – 0.9); $p=0.034$ |
| Based in an acute general hospital | - | 1.8 (1.1 – 2.8); $p=0.016$ | - |
| Working in inpatient wards / ED / ITU | - | - | 1.4 (1.1 – 1.8); $p=0.006$ |
| Reported adequate PPE at work | 0.7 (0.5 – 0.8); $p<0.001$ | 0.7 (0.6 – 0.9); $p=0.001$ | 0.7 (0.5 – 0.8); $p<0.001$ |
| Wellbeing support available at work | 0.6 (0.5-0.7); $p<0.001$ | 0.5 (0.4 – 0.7); $p<0.001$ | 0.5 (0.4 – 0.7); $p<0.001$ |
| Denied morally uncomfortable changes | 0.5 (0.4 – 0.6); $p<0.001$ | 0.6 (0.5 – 0.7); $p<0.001$ | 0.4 (0.3 – 0.5); $p<0.001$ |
| Redeployed | - | - | 1.5 (1.2 – 1.9); $p<0.001$ |
| Increased working hours | - | - | 1.3 (1.1 – 1.6); $p=0.006$ |
| Admitted to hospital with Covid-19* | 1.4 (1.1-1.7); $p=0.009$ | - | 1.7 (1.3 – 2.2); $p<0.001$ |

#### ii. Depressive symptoms

Smokers were 50% more likely to report depressive symptoms (OR 1.5 [1.1-1.9]; *p*=0.005). A history of mental health conditions had the strongest association, with two and half times greater odds of hospital HCW reporting depressive symptoms (OR 2.5 [2.1-3.0]; *p*<0.01). There was an almost two-fold increase in odds of depressive symptoms when the participant was based in an acute general hospital compared to a mental health setting (OR 1.8 [1.1-2.8]; *p*=0.016). On the other hand, alcohol consumers were 20% less likely to experience these symptoms (OR 0.8 [0.7-0.9]; *p*=0.028). Furthermore, staff with adequate PPE availability, adequate wellbeing support and those who did not report morally uncomfortable changes in the way they worked were up to 50% less likely to report depressive symptoms (*p*≤0.001; see Table 3 and supplementary Fig. 3). There were no significant associations between the remaining exposures and depressive symptoms (supplementary Table 2).

#### iii. Symptoms of post-traumatic stress disorder

A history of mental health conditions was associated with a two-fold increased odds of clinically significant PTSD symptoms (OR 2.1 [1.7-2.5]; *p*<0.001). Several exposures were associated with increased likelihood of clinically significant PTSD symptoms: female gender, history of physical illness, smoking, being based in inpatient wards, ED or ITU, redeployment, and admission to hospital in self, close family or friend with Covid-19 (*p*<0.05; see Table 3). The exposures of alcohol consumption and working as a doctor or a nurse were associated with a 20% lower likelihood of reporting clinically significant PTSD symptoms (*p*<0.05; see Table 3). There were 30-50% less odds of clinically significant PTSD symptoms when there were adequate PPE and wellbeing support available (*p*<0.001; see Table 3 and supplementary Fig. 4). Participants who reported that no morally uncomfortable changes took place in the way they worked demonstrated approximately 60% less odds of clinically significant PTSD symptoms (OR 0.4 [0.3-0.5]; *p*<0.001). There were no significant associations between the remaining exposures and PTSD symptoms (supplementary Table 2).

## Discussion

We found that around a third of hospital HCW reported clinically significant symptoms of anxiety and depression. A quarter reported clinically significant PTSD symptoms. Previous studies reported a baseline prevalence of clinically significant PTSD symptoms (also defined as an IES-R score ≥33) amongst 15-16% of HCW from the UK.^24,25^ No comparable published data for UK HCW PHQ-4 scores were found. However, the prevalence rate of anxiety and depression amongst the UK general population was previously reported to be around 19.7%.^26^ The higher rate of PTSD symptoms identified in the current study might be associated with working in a hospital setting during the Covid-19 pandemic. Nevertheless, the prevalence of symptoms of anxiety and depression amongst HCW in this study was lower compared to the data from China.^7^ Differences could be attributed to a multitude of factors including cultural, political and socio-economic variations across the two study populations.

In adjusted analysis a previous history of mental health conditions consistently demonstrated odds of greater than two in participants reporting clinically significant symptoms of anxiety, depression and PTSD. In contrast, the availability of adequate PPE, access to wellbeing support and not experiencing morally uncomfortable changes in the way they worked were significantly negatively associated with participants reporting these symptoms. Younger participants aged 40 or under demonstrated greater odds of reporting clinically significant anxiety symptoms. Smoking was associated with depression and PTSD symptoms but not anxiety. Female gender and a hospital admission in self, a close family member or a friend were associated with anxiety and PTSD symptoms but not depression. Several other exposures were associated with PTSD symptoms but not anxiety or depression (e.g. redeployment, increased working hours and working in inpatient wards, ED or ITU).

Our findings may prompt healthcare employers to focus their attention on the provision of specific interventions that may protect HCW against adverse mental health impacts during crises such as the Covid-19 pandemic. These may include the provision of greater access to wellbeing support for staff, ensuring the availability of adequate PPE and protection from exposure to moral dilemmas in the workplace. Furthermore, careful workforce planning to mitigate the adverse effects of redeployment and minimising the risk of viral infection may reduce the risk of staff experiencing PTSD symptoms. Attention should also be given to staff at greater risk such as those with a history of mental health conditions, female staff and smokers. Special consideration and additional support in the workplace could also be considered for younger employees, redeployed staff members and those working in potentially high-risk areas (e.g. inpatient wards, ED and ITU)

There were some unexpected findings in this analysis. One such finding was the protective effect of working as a doctor or a nurse had on participants reporting clinically significant symptoms of anxiety and PTSD. This could be attributed to factors such as training, experience and coping mechanisms or resilience from previous working practices in stressful healthcare environments. Furthermore, there was no statistically significant increase in odds of self-reported mental health symptoms amongst staff undertaking patient-facing duties compared to staff in other roles. Therefore, it is important to ensure support is available to staff in all job roles who may potentially be at risk and not just frontline clinical staff. We also observed reduced odds of clinically significant symptoms of depression and PTSD reported by participants who consumed any amount of alcohol. The clinical significance and relevance of the latter association are unknown.

There were some limitations in our study. The time elapsed between traumatic exposure and the onset of symptoms is key to making a diagnosis of PTSD. However, the aim of the study was not to make diagnoses of mental health disorders but to screen the target population for the presence of clinically concerning symptoms. The survey was conducted relatively close to the duration of the UK’s Covid-19 pandemic peak. The elevated scores on the IES-R may be representative of an acute stress reaction which usually resolves within a few months. Further follow-up of these participants is required to ascertain the persistence of symptoms—a planned analysis by our study group. Our data is from a cross-sectional survey. Therefore, causal inferences cannot be made. Furthermore, the data were collected through a self-report questionnaire which is at risk of responder bias. There were also several strengths. This study is one of the first in the UK to report on the mental health impact of working during the Covid-19 pandemic on hospital staff. These findings may be generalisable to the wider UK population of hospital employees given the relatively large sample size and representative demographic sample (supplementary Table 3).

During the Covid-19 pandemic, there were high prevalence rates of common mental health symptoms in hospital HCW in the UK, especially amongst those with a past history of mental health conditions. Occupational interventions such as adequate PPE and wellbeing support availability, and reduced exposure to moral dilemmas appear to protect hospital HCW against these symptoms.

## Data Availability

All relevant data and results included in this article have been published along with the article and its supplementary information files. Anonymised data can be obtained on reasonable request from the corresponding author at the end of the STAT-STRESS Covid-19 study.

## Acknowledgements

We would like to acknowledge our study sponsor the Birmingham Women’s and Children’s Hospital NHS Foundation Trust. We are grateful for the support from the research and development teams and executive leadership of all ten acute National Health Service hospital Trusts in the West Midlands, UK. We would further like to thank the West Midlands Research Collaborative and West Midlands Surgical Society for their endorsement of this study. We extend our gratitude to Mr S Karandikar (Heartlands Hospital), Mr R.M. Tirumularaju (Good Hope Hospital), Dr E Plunkett (Queen Elizabeth Hospital Birmingham) and Dr E Jenkinson (New Cross Hospital) from the West Midlands, UK for their assistance with participant recruitment. Finally, we would like to acknowledge and thank all our participants for their time spent taking part in this study.

## Author contributions

KW and PP designed the study, created the survey, analysed the data and wrote the main manuscript. DN and KI were involved in data interpretation and co-writing the main manuscript. JLF was involved in survey design and participant recruitment. PL, CVT, HB and ADB were involved in study promotion, participant recruitment, reviewing and editing of the manuscript. TI was the most senior author. He provided supervision and guidance for the study. He was involved in study design, management, data interpretation, reviewing and editing the final manuscript.

## Statistical analysis

Statistical analysis was performed by Dr Kasun Wanigasooriya. The statistical analysis was overseen by Mr Peter Nightingale, Statistician at the University Hospitals Birmingham NHS Foundation Trust, UK.

## Funding

The University Hospitals Birmingham Charity covered the operational expenses incurred during this study (Remittance Reference No: C05976).

## Declaration of interest

None.

## Notes

### Competing Interest Statement

The authors have declared no competing interest.

### Author Declarations

The study was approved by the UK Health Research Authority (HRA, Reference: 20/HRA/2865). Research Ethics Committee approval was not required for this study and this was confirmed by the HRA. Site specific approval was obtained for participant recruitment from each of the research and development departments of all participating hospital Trusts (University Hospitals Birmingham NHS Foundation Trust, Sandwell and West Birmingham Hospitals NHS Trust, Birmingham Women's and Children's Hospital NHS Trust, Royal Wolverhampton Hospital NHS Trust, Walsall Healthcare NHS Trust, Dudley Group of Hospitals NHS Trust, University Hospital Coventry and Warwickshire NHS Trust, Coventry and Warwickshire Partnership NHS Trust, Black Country Mental Health Partnership Trust, Birmingham and Solihull Mental Health NHS Trust). Informed consent was obtained from all participants and recorded electronically at the start of the study.

