## supplementary Fig. 1 for "Mental health symptoms in a cohort of hospital healthcare workers following the first peak of the Covid-19 pandemic in the United Kingdom"

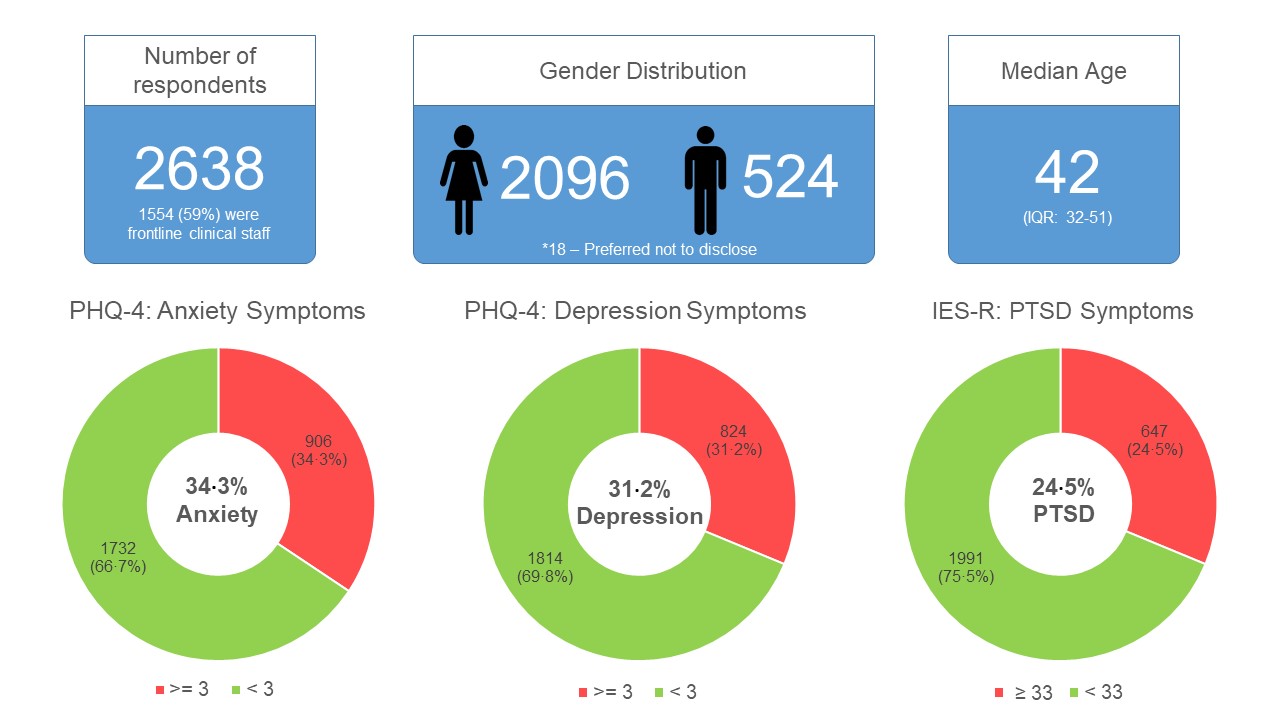

**Supplementary Fig. 1:** A summary of the key demographic data of survey participants, PHQ-4 and IES-R scores
