## supplementary Fig. 2 for "Mental health symptoms in a cohort of hospital healthcare workers following the first peak of the Covid-19 pandemic in the United Kingdom"

**PHQ-4: Anxiety Subscale Score >= 3**

**Odds Ratio (95% CI)**

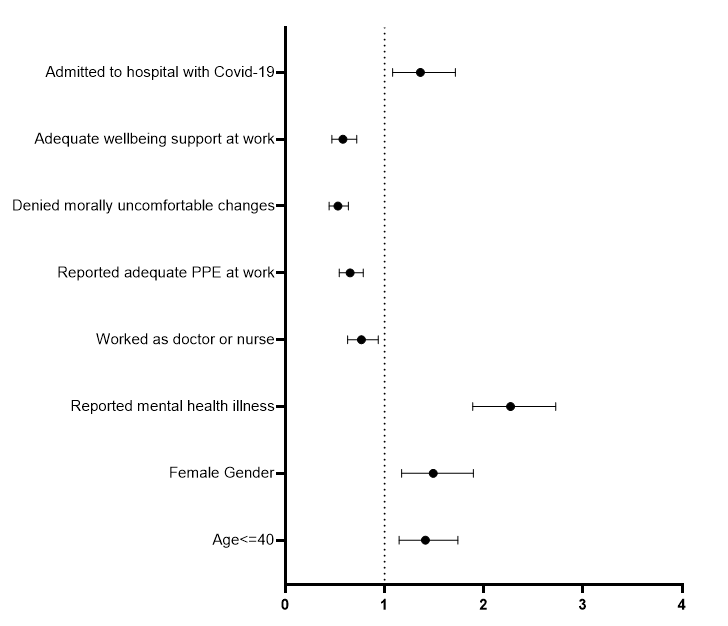

| Admitted to hospital with Covid-19 | 1·4 (1·1-1·7); p*=*0·009 |
| --- | --- |
| Wellbeing support available at work | 0·6 (0·5-0·7); p*<*0·001 |
| Denied morally uncomfortable changes | 0·5 (0·4 – 0·6); p*<*0·001 |
| Reported adequate PPE at work | 0·7 (0·5 – 0·8); p*<*0·001 |
| Worked as a doctor or nurse | 0·8 (0·6-0·9); p*=*0·009 |
| A history of mental health conditions | 2·3 (1·9 – 2·7); p*<*0·001 |
| Female gender | 1·5 (1·2 – 1·9); p*=*0·001 |
| Age <= 40 | 1·4 (1·1 – 1·7); p*=*0·001 |

**Odds Ratio (95% CI)**

**Supplementary Figure 2:** Factors associated with clinically significant anxiety symptoms in adjusted analysis
