## supplementary Fig. 3 for "Mental health symptoms in a cohort of hospital healthcare workers following the first peak of the Covid-19 pandemic in the United Kingdom"

**PHQ-4: Depression Subscale Score >= 3**

**Odds Ratio (95% CI)**

| Wellbeing support available at work | 0·5 (0·4 – 0·7); p*<*0·001 |
| --- | --- |
| Denied morally uncomfortable changes | 0·6 (0·5 – 0·7); p*<*0·001 |
| Reported adequate PPE at work | 0·7 (0·6 – 0·9); p*=*0·001 |
| 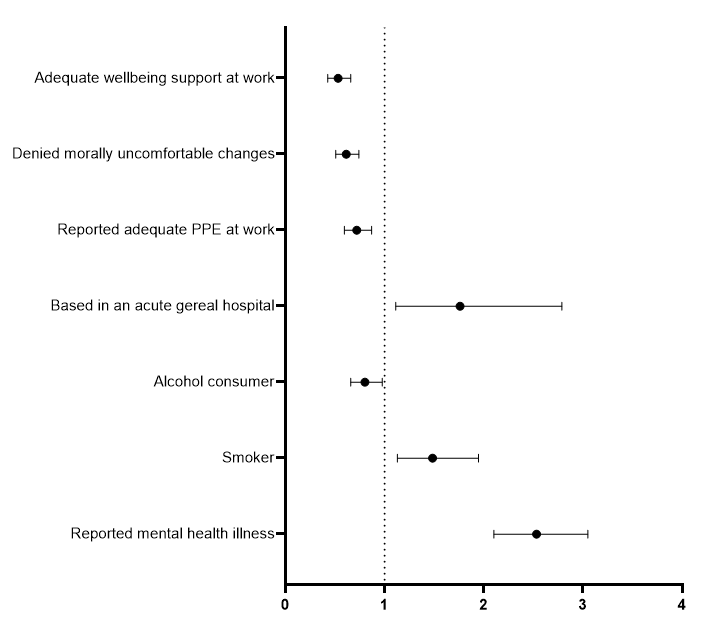 Based in an acute general hospital | 1·8 (1·1 – 2·8); p*=*0·016 |
| Alcohol consumer | 0·8 (0·7-0·9); p*=*0·028 |
| Smoker | 1·5 (1·1 – 1·9); p*=*0·005 |
| A history of mental health conditions | 2·5 (2·1 – 3·0); p*<*0·001 |

**Odds Ratio (95% CI)**

**Supplementary Figure 3:** Factors associated with clinically significant depressive symptoms in adjusted analysis
