## supplementary Fig. 4 for "Mental health symptoms in a cohort of hospital healthcare workers following the first peak of the Covid-19 pandemic in the United Kingdom"

**Clinically Significant PTSD Symptoms: IES-R Score >= 33**

**Odds Ratio (95% CI)**

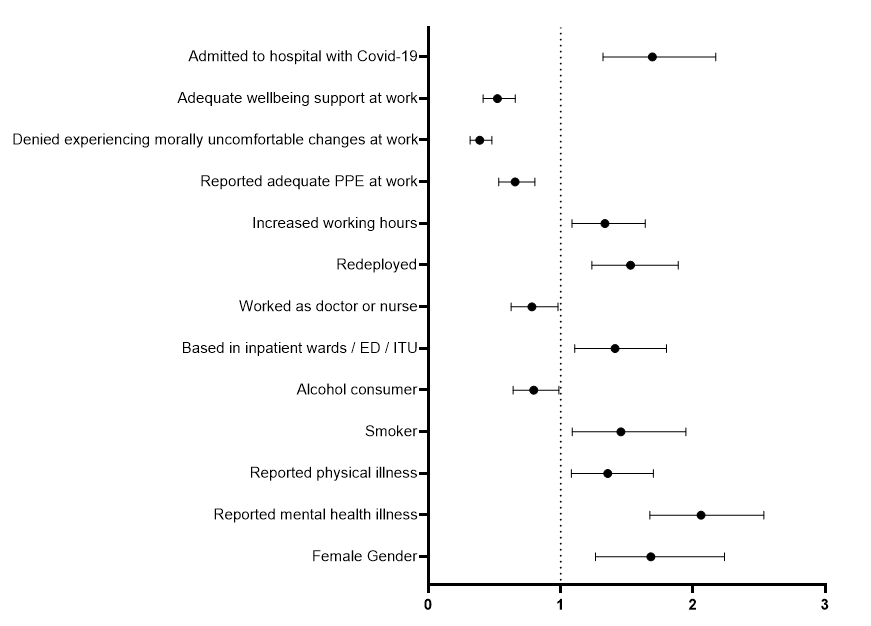

| Admitted to hospital with Covid-19* | 1·7 (1·3 – 2·2); p*<*0·001 |
| --- | --- |
| Wellbeing support available at work | 0·5 (0·4 – 0·7); p*<*0·001 |
| Denied morally uncomfortable changes | 0·4 (0·3 – 0·5); p*<*0·001 |
| Reported adequate PPE at work | 0·7 (0·5 – 0·8); p*<*0·001 |
| Increased working hours | 1·3 (1·1 – 1·6); p*=*0·006 |
| Redeployed | 1·5 (1·2 – 1·9); p*<*0·001 |
| Worked as a doctor or nurse | 0·8 (0·6 – 0·9); p*=*0·034 |
| Working in inpatient wards / ED / ITU | 1·4 (1·1 – 1·8); p*=*0·006 |
| Alcohol consumer | 0·8 (0·6 – 0·9); p*=*0·038 |
| Smoker | 1·5 (1·1 – 1·9); p*=*0·012 |
| A history of physical illness | 1·4 (1·1 – 1·7); p*=*0·009 |
| A history of mental health conditions | 2·1 (1·7 – 2·5); p*<*0·001 |
| Female gender | 1·7 (1·3 – 2·2); p*<*0·001 |

**Odds Ratio (95% CI)**

**Supplementary Figure 4:** Factors associated with clinically significant symptoms of PTSD in adjusted analysis
