## supplementary Table 1 for "Mental health symptoms in a cohort of hospital healthcare workers following the first peak of the Covid-19 pandemic in the United Kingdom"

|  | N | % |
| --- | --- | --- |
| *Where staff were based within the hospital during the pandemic peak* |  |  |
| *All inpatient wards* | 704 | 26·7% |
| *Non patient areas (e.g. administrative/office space, labs, kitchen, switchboard, stores, plant rooms)* | 507 | 19·2% |
| *Multiple locations in the hospital* | 446 | 16·9% |
| *Critical Care / High Dependency Unit* | 382 | 14·5% |
| *Outpatients / Clinic rooms* | 226 | 8·6% |
| *Emergency Department* | 123 | 4·7% |
| *Theatre / procedure rooms (e.g. endoscopy)* | 121 | 4·6% |
| *Radiology* | 83 | 3·1% |
| *Pharmacy* | 35 | 1·3% |
| *Mortuary* | 10 | 0·4% |
| *Restaurant, café or shop* | 1 | >0·1% |
| *Staff job titles* |  |  |
| *All nurses* | 775 | 29·4% |
| *All doctors* | 460 | 17·4% |
| *Healthcare Assistant / Support Worker / Nursing Associate* | 228 | 8·6% |
| All managers | 147 | 5·6% |
| *Secretarial staff/Medical Secretary/Personal Assistant* | 124 | 4·7% |
| *Healthcare Records Clerk / Ward Clerk / Other admin* | 123 | 4·7% |
| *Healthcare Scientist (e.g. biochemist)* | 94 | 3·6% |
| *Physiotherapist* | 93 | 3·5% |
| *Operating Department Practitioner (ODP)* | 68 | 2·6% |
| *Radiographer* | 58 | 2·2% |
| *Corporate Services (e.g. finance/HR/executives/directors)* | 56 | 2·1% |
| *Midwife* | 56 | 2·1% |
| *Receptionist* | 53 | 2·0% |
| *Healthcare Technician* | 37 | 1·4% |
| *Occupational Therapist* | 34 | 1·3% |
| *Other Pharmacy Roles* | 33 | 1·3% |
| *Pharmacist* | 31 | 1·2% |
| *Medical Associate Professional (e.g. Physicians Associates)* | 27 | 1·0% |
| *Dietician* | 18 | 0·7% |
| *Estate services* | 18 | 0·7% |
| *IT Staff* | 13 | 0·5% |
| *Phlebotomist* | 13 | 0·5% |
| *Speech and Language Therapist* | 13 | 0·5% |
| *Optician/Orthoptist* | 12 | 0·5% |
| *Domestic (e.g. catering, housekeeping)* | 10 | 0·4% |
| *Psychologist* | 10 | 0·4% |
| *Student* | 9 | 0·3% |
| *Porter* | 8 | 0·3% |
| *Chaplaincy* | 7 | 0·3% |
| *Podiatrist* | 3 | 0·1% |
| *Switchboard Operator* | 3 | 0·1% |
| *Volunteer* | 2 | 0·1% |
| *Allied Health Professional* | 1 | >0·1% |
| *Prosthetist / Orthotist* | 1 | >0·1% |

**Supplementary Table 1:** Staff base-locations in the workplace and their job titles
