## supplementary Table 2 for "Mental health symptoms in a cohort of hospital healthcare workers following the first peak of the Covid-19 pandemic in the United Kingdom"

|  | **PHQ-4 Anxiety Subscale**  **Score** ≥ **3** | | **PHQ-4 Depression Subscale**  **Score** ≥ **3** | | **PTSD Symptoms**  **IES-R Score** ≥ **33** | |
| --- | --- | --- | --- | --- | --- | --- |
|  | Univariate  Analysis % | Logistic Regression  OR (95% CI) | Univariate  Analysis % | Logistic Regression  OR (95% CI) | Univariate  Analysis % | Logistic Regression  OR (95% CI) |
| 1. *Denied morally uncomfortable changes in the way they worked (moral dilemmas)* | 25·5% vs· 43·7%  p<0·001 | 0·529 (0·440-0·635)  p<0·001 | 24·9% vs· 37·8%  p < 0·001 | 0·614  (0·508-0·742)  p < 0·001 | 14·3% vs· 35·3%  p < 0·001 | 0·389  (0·315–0·481)  p < 0·001 |
| 2. *Wellbeing support available at work* | 30·7% vs· 47·4%  p < 0·001 | 0·581  (0·470-0·719)  p < 0·001 | 27·4% vs· 45·1%  p < 0·001 | 0·531  (0·428–0·659)  p < 0·001 | 20·6% vs· 38·7%  p <0·001 | 0·522  (0·444–0·657)  p < 0·001 |
| 3. *Adequate PPE availability at work* | 28·0% vs· 42·2%  p < 0·001 | 0·653  (0·544-0·785)  p < 0·001 | 25·6% vs· 38·2%  p < 0·001 | 0·720  (0·596–0·869)  p = 0·001 | 18·7% vs· 31·6%  p < 0·001 | 0·655  (0·523–0·806)  p < 0·001 |
| 4. *Alcohol consumer* | 32·5% vs· 37·8%  p = 0·007 | NS | 28·6% vs· 36·1%  p < 0·001 | 0·802  (0·659-0·977)  p = 0·028 | 21·9% vs· 19·211  p < 0·001 | 0·796  (0·641 – 0·987)  p = 0·038 |
| 5. *Working as a doctor or a nurse* | 30·8% vs· 37·5%  p < 0·001 | 0·766  (0·627-0·936)  p = 0·009 | 34·7% vs· 32·9%  p < 0·001 | NS | NS | 0·783  (0·625–0·981)  p = 0·034 |
| 6. *Being in a relationship* | NS | NS | 29·8% vs· 35·9%  p =0·004 | NS | 23·3% vs· 28·5%  p = 0·009 | NS |
| 7. *Having dependents at home* | 32·4% vs· 36·3%  p=0·034 | NS | 28·9% vs· 33·5%  p = 0·011 | NS | NS | NS |
| 8. *History of mental health conditions* | 46·5% vs· 26·3%  p < 0·001 | 2·269  (1·889–2·725) p < 0·001 | 44·8% vs· 22·3%  p < 0·001 | 2·531  (2·101–3·049)  p < 0·001 | 40·0% vs· 18·5%  p < 0·001 | 2·061  (1·674–2·536)  p < 0·001 |
| 9. *Female gender* | 36·4% vs· 25·6%  p < 0·001 | 1·490  (1·172–1·895)  p = 0·001 | 32·7% vs· 24·8%  p < 0·001 | NS | 26·4% vs· 16·0%  p < 0·001 | 1·682  (1·264–2·239)  p < 0·001 |
| 10. *Smoker* | 43·5% vs 33·2%  p < 0·001 | NS | 43·9% vs· 29·6%  p < 0·001 | 1·483  (1·130-1·947)  p = 0·005 | 35·2% vs· 23·1%  p < 0·001 | 1·455  (1·087 – 1·946) p = 0·012 |
| 11. *Age ≤ 40* | 39·1% vs· 30·1%  p < 0·001 | 1·412  (1·146–1·739)  p = 0·001 | 35·4% vs 27·5%  p < 0·001 | NS | 26·9% vs· 22·4%  p = 0·008 | NS |
| 12. *A hospital admission with Covid-19 (self, close family, or friend)** | 41·8% vs· 32·8%  p < 0·001 | 1·361  (1·080-1·714)  p = 0·009 | NS | NS | 35·6% vs· 22·2%  p < 0·001 | 1·693  (1·320–2·172)  p < 0·001 |
| 13. *Redeployment* | NS | NS | NS | NS | 31·4% vs· 21·1%  p <0·001 | 1·528  (1·236–1·889)  p < 0·001 |
| 14. *Increased working hours* | NS | NS | NS | NS | 26·9% vs· 23·0%  p = 0·025 | 1·334  (1·086–1·639)  p = 0·006 |
| 15. *Based in inpatient wards / ED / ITU* | NS | NS | NS | NS | 29·4% vs· 20·4%  p < 0·001 | 1·411  (1·106 – 1·801)  p = 0·006 |
| 16. *History of physical illness* | 37·7% vs· 32·7%  p = 0·018 | NS | 36·6% vs· 29·0%  p < 0·001 | NS | 29·9% vs· 22·3%  p < 0·001 | 1·356  (1·081–1·701)  p = 0·009 |
| 17. *Getting Covid-19* | NS | NS | NS | NS | 29·0% vs· 22·8%  p = 0·001 | NS |
| 18. *Working in healthcare for 10 years or less* | 38·2% vs· 32·2%  p = 0·002 | NS | 37·2% vs· 27·9%  p < 0·001 | NS | 28·7% vs· 22·2%  p < 0·001 | NS |
| 19. *Based at an acute general hospital* | NS | NS | NS | 1·760  (1·112–2·786)  p = 0·016 | NS | NS |
| 20. *Staff undertaking patient-facing roles* | NS | NS | NS | NS | 27·0% vs· 20·9%  p < 0·001 | NS |
| 21. *Being a staff member from BAME group* | NS | NS | NS | NS | 28·4% vs· 23·7%  p = 0·037 | NS |
| 22. *Staff who have emigrated to the UK as adults* | NS | NS | NS | NS | NS | NS |
| 23. *Working in a hospital that treated Covid-19 patients* | NS | NS | NS | NS | NS | NS |
| 24. *Cohabitant getting Covid-19* | NS | NS | NS | NS | NS | NS |

**Supplementary Table 2:** A visual summary of all significant and non-significant results from the univariate univariate analysis and adjusted logistic regression analysis

NS = Not statistically significant	Statistically significant result

*Close family member – nuclear family, first degree relative whom the respondent lived with or associated on at least once a week; a close friend: A friend whom the participant lived with or associated with at least once a week.
