## supplementary Table 3 for "Mental health symptoms in a cohort of hospital healthcare workers following the first peak of the Covid-19 pandemic in the United Kingdom"

|  | Hospital Trust A | Hospital Trust B | Hospital Trust C | Our sample |
| --- | --- | --- | --- | --- |
| *Total staff* | 19942 | 5666 | 9590 | 2638 |
| *Gender*  *Female*  *Male*  *Prefer not to say* | 14991 (75·2%)  4951 (24·8%)  .. | 4652 (82·1%)  1014 (17·9%)  .. | 7557 (78·8%)  2033 (21·2%)  .. | 1983 (75·2%)  637 (24·1%)  18 (0·7%) |
| *Ethnicity*  *BAME*  *Caucasian*  *Not stated* | 6510 (32·6%)  12,711 (63·7%)  721 (3·6%) | 1279 (22·6%)  3533 (62·4%)  854 (15·1%) | 2858 (29·8%)  6592 (68·7%)  140 (1·5%) | 455 (17·2%)  2183 (82·8%)  .. |
| *Job title:*  Medical and Dental  Nursing and Midwifery  Other | 2397 (12·0%)  5572 (27·9%)  11973 (60·1%) | 612 (10·8%)  1866 (32·9%)  3188 (56·3%) | 1032 (10·8%)  2679 (27·9%)  5879 (61·3%) | 460 (17·4%)  831 (31·5%)  1347 (51·1%) |

**Supplementary Table 3:** Baseline demographic data from some of the hospitals in the county
